# Chest Compression Synchronized Ventilation (CCSV) during cardiac arrest in animals and humans: A scoping review

**DOI:** 10.1101/2025.05.28.25328471

**Authors:** Roman Brock, Christoph Veigl, Andrea Kornfehl, Johannes Wittig, Sabine Heider, Karina Tapinova, Erwin Snijders, Sabine Dunkl, Daniel Grassmann, Birgit Heller, Mario Krammel, Sebastian Schnaubelt

## Abstract

**Background:** Ventilation during cardiopulmonary resuscitation (CPR) should be effective. Chest Compression Synchronized Ventilation (CCSV) is a novel approach aimed at optimizing gas exchange and hemodynamics by synchronizing mechanical ventilation with chest compressions. However, its clinical value, safety profile, and implementation barriers remain unclear. We thus aimed to systematically synthesize existing evidence on the use of CCSV during cardiac arrest in animals and humans.

**Methods:** We conducted a scoping review and systematically searched five databases (Medline, Embase, CENTRAL, Scopus, Web of Science) up to May 2025. Studies investigating CCSV or mechanistically related ventilation strategies during cardiac arrest were included regardless of study design, language, or publication date. Data were charted for study characteristics, outcomes, and adverse events.

**Results:** Thirty-one studies published between 1980 and 2025 were included. Most were animal studies (n=19), primarily conducted in pigs, with limited human data (n=9). CCSV showed favorable effects on arterial oxygenation, carbon dioxide clearance, and hemodynamic parameters. Some studies reported improved return of spontaneous circulation and cerebral oxygenation compared to conventional ventilation modes. Adverse events such as pneumothorax and lung injury were inconsistently reported and mostly limited to animal models.

**Conclusions:** Available data on CCSV suggests potential physiological benefits during CPR, particularly in experimental settings. Human data remain scarce, and larger, prospective human trials are essential to evaluate clinical effectiveness, guide implementation, and assess risks compared to conventional ventilation strategies.

## Background

Current European Resuscitation Council (ERC) guidelines on cardiopulmonary resuscitation (CPR) recommend uninterrupted chest compressions with asynchronous ventilation with a rate of 10 breaths per minute following advanced airway management during advanced life support (ALS). However, recommendations regarding ventilation parameters, the use of mechanical ventilators and specific ventilation modes during CPR remain scarce. ^1^ American Heart Association guidelines recommend tidal volumes between 500-600ml delivered with a rate of 10/min if advanced airway control has been performed. However, the use of simultaneous compression and ventilation is discouraged ^2^ based on a study by *Krischer et al*. However, this study has to be viewed with caution in this context as it describes chest compression at a rate of 40/min with synchronized ventilation with 80 mmHg of airway pressure (SC-V) ^3^ Regarding mechanical ventilation during CPR, there is no unanimous opinion: In an opinion survey in 54 countries, *Cordioli et al.* report that the most common ventilation mode is volume-controlled ventilation, followed by pressure-controlled ventilation and continuous positive pressure ventilation. The fraction of inspired oxygen (FiO_2_) was set to 100% or titrated to achieve a target of 94% of peripheral oxygen saturation (SpO_2_) most of the time.^4^ Various ventilations modes or variations thereof are described during CPR. ^5^ For the present review we used the following definitions of common ventilation modes:

- Intermittent positive pressure ventilation (IPPV): IPPV describes a conventional volume-controlled ventilation mode. A set tidal volume is applied with a set ventilation rate. Additionally, the application of positive end-expiratory pressure (PEEP) is possible. Maximum airway pressure can be limited by the ventilator.^6,7^
- Bi-Level Positive Airway Pressure Ventilation (BiPAP): BiPAP ventilation is characterized by two different, predefined pressure levels (P_low_ and P_high_), cycled at ventilation rate. It allows for pressure-controlled ventilation while simultaneously permitting spontaneous breathing with pressure support in any ventilation phase. Synchronization of mandatory and spontaneous ventilation is regulated by the ventilator. This mode is available in a variety of commercial ventilators. However, nomenclature might differ between companies.^8^
- Chest Compression Synchronized Ventilation (CCSV, WEINMANN, Emergency Medical Technology GmbH + Co. KG, Hamburg, Germany): CCSV is a ventilation mode specifically designed for CPR. Expiratory airflow caused by chest compression is used as a signal to trigger pressure-controlled mechanical ventilation at 40 mbar or 60 mbar immediately. Therefore, air is pushed inside the patient simultaneously to chest compression (with the same rate). PEEP can be necessary for correct triggering. This mechanism is proposed to result in increased intrathoracic pressure concentrated at the heart and in further consequence positive hemodynamic effects as increased mean arterial pressure. However, data in humans is currently limited. ^9^ The idea of ventilation applied simultaneously with chest compressions was first introduced in 1980 by Chandra et al. when they explored a so called “new CPR” (40 compressions per minute, simultaneous ventilation with 60-110 cmH_2_O airway pressure) in eleven patients after conventional means of resuscitation had failed, and reported higher carotid flow and pressure in the radial artery.^10^ In this context, the study of Krischer et al. has to be referenced, evaluating a synchronized compression-ventilation technique at a rate of 40/min with at 80 mmHg of airway pressure resulting in a 11% lower survival rate in non-traumatic cardiac arrests. ^3^

Regarding the superiority of manual or mechanical ventilation during CPR, literature is conflicting: CPR results in oscillations in airway pressures and can cause reversed air flow during inspiratory and expiratory ventilation phases.^11^ This can cause deviations from parameters set on the ventilator^12^ and problems due to trigger related issues. ^13^ Whatever the ventilation technique is, a measurement of ventilation parameters as a form of feedback is recommended^14^ to ensure ventilation quality, which may be more challenging during manual ventilation because additional devices are needed. A recent clinical study on CCSV suggest that the intervention may be effective in maintaining gas exchange during CPR ^15^ and the mode seems to be in use internationally in the absence of international recommendations for its use.

A comprehensive overview of published literature on CCSV, and thus a descriptive summary of the current knowledge base along with relevant gaps for future research is lacking. We therefore conducted a scoping review to provide an overview of clinical and pre-clinical studies of CCSV during cardiac arrest.

## Methods

This scoping review collected publications on CCSV during cardiac arrest in animals and humans. Slightly different but mechanistically similar algorithms were also eligible for the review (Cardiopulmonary Ventilation (CPV), Simultaneous Compression Ventilation (SC-V), automatic compression synchronous ventilation (ACSV)). Our protocol was designed according to the PRISMA Extension for Scoping Reviews (PRISMA-ScR),^16^ following these key features:

- Sample: Humans and animals during cardiac arrest
- Phenomenon of Interest: Ventilation with CCSV during cardiac arrest
- Design: Structured literature research with predefined search strategy
- Evaluation:

◦ Patient centered outcomes: Return of spontaneous circulation (ROSC), survival to hospital discharge, 30-days survival, neurological outcome
◦ Changes in vital signs and laboratory measurements: Partial pressures of oxygen (PO_2_), carbon dioxide (PCO_2_), pH, blood pressures, peripheral oxygen saturation (SpO_2_), regional tissue oxygenation (rSO_2_), biomarkers of neuroinflammation
◦ Adverse events: Ventilator associated lung injury (VALI), pneumothorax, histological changes in lung tissue.
◦ Status of CCSV in current research and practice: Barriers and facilitators towards use, positive and negative perceptions
- Research Type: All literature published in peer-reviewed journals was eligible for inclusion. All years and all languages were included as long as there was an English abstract. There was no restriction of publications regarding time of publication.

### Search strategy and evidence selection

We performed a systematic search of Medline, Embase, Central, Web of Science and Scopus databases on March 3^rd^ 2025, with an update on May 25^th^ 2025. The search strategy was designed by an information specialist *(BH)* and further refined within the research team. The full strategy for all databases can be found in the Supplements.

Search results were analyzed for eligibility in a two-step process: First, duplicates were removed, and titles and abstracts were evaluated by two independent reviewers. Conflicts were resolved by consensus. Secondly, the full texts of eligible sources were retrieved and assessed in the same multi-reviewer process. Excluded sources were documented with justification. Additionally, references of included sources were searched for relevant publications and screened in the same manner. Rayyan (Rayyan.ai) and Zotero 7.0.7 (Corporation for Digital Scholarship) were used for evidence selection.

### Data extraction and analysis

Data extraction was performed on the included full text results. Beside study type, population size, aims and interventions/comparisons, the articles were evaluated for the evaluation features described above (patient centered outcomes, changes in vitals and lab values, adverse events). Descriptive statistics were performed with R 4.4.1 (R Foundation for Statistical Computing, Vienna, Austria).

## Results

### Screening

The structured search yielded a total of 1,292 results. After removal of duplicates, 753 records were screened for eligibility, resulting in 50 reports for further examination. Nineteen records were excluded due to lack of relation to the topic of interest. Additionally, in three cases, abstracts and their associated articles published afterwards were found independently resulting in the exclusion of the abstract. One abstract was published twice and therefore excluded once. In three cases, the full-texts could not be retrieved, limiting data extraction to abstracts in these cases. ^17–19^ Seven additional reports were identified by manual research. Of these, five were included in the review after full-text screening. In total, 32 studies were included in the review *(Figure 1)*.

**Figure 1:**
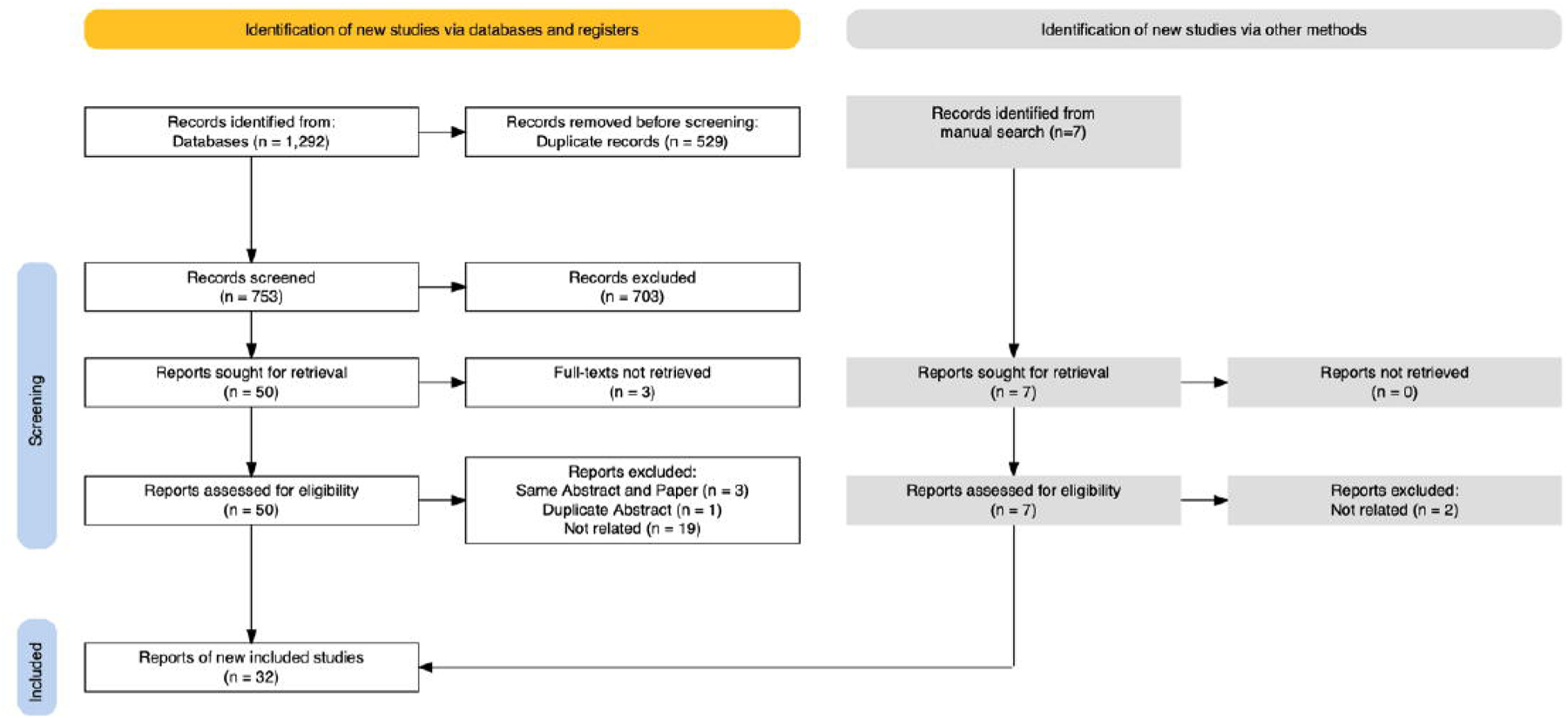
Flow diagram of the screening and inclusion I exclusion process.

### Characteristics of selected sources

A total of 32 reports published between 1980 and 2025 were included in the review and further assessed: Twenty-three reports (72%) focused on CCSV directly, seven reports (22%) described other ventilation modes or experimental ventilation protocols during chest compression^3,10,17,20–23^, and two (6%) explored protocols for ventilation between compressions.^24,25^ Most reports originated from Germany (n=15, 47%), USA (n=4, 13%) or China (n=3, 9%). The majority of studies were designed as randomized controlled trials (n=13, 41%) ^3,15,17–19,25–32^ and conducted in animals (pigs; n=18, 58%) ^17,21,22,25–39^. Nine studies (28%) report outcomes in humans ^3,10,15,19,23,24,40–42^, one (3%) in human cadavers^18^, and another one (3%) was conducted on manikins.^43^ Of note, one case report^40^, one editorial^44^, and one review^44^ resulted from the database search. To estimate the resource setting the reports originated from, the respective countries have been categorized according to the World Bank classifications^45^ *(see Table 1 for details)*.

**Table 1:**
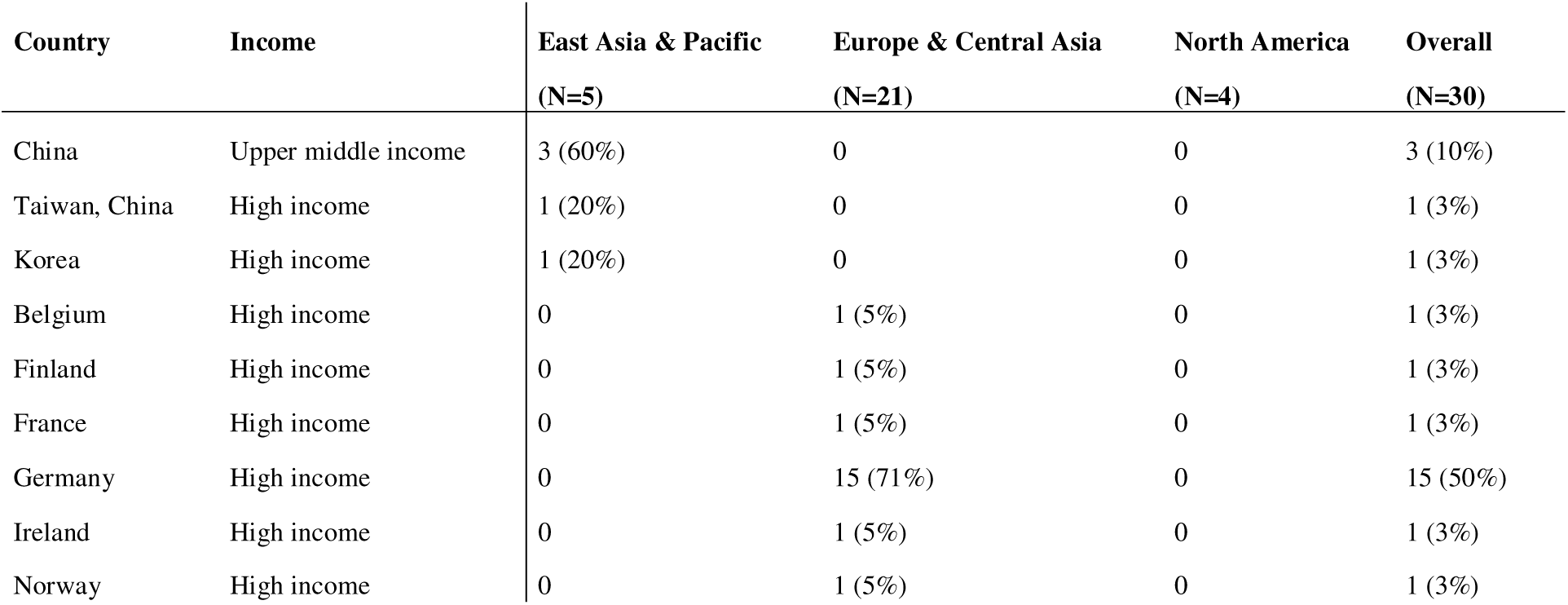
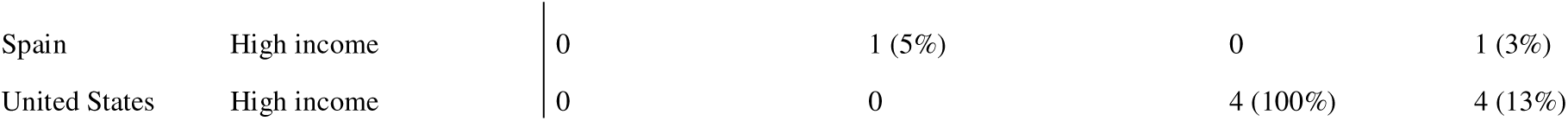
Income classification table according to the World Bank classification^45^. In two cases, the country could not be determined due to missing full-text.

### Outcome parameters

Fourteen reports (44%)^15,17,19–21,27,28,30–32,34,37,38,42^ included blood gas analyses results (arterial pO_2_ and pCO_2_ in all fourteen cases, and venous pO_2_ and pCO_2_ in only three cases^30,34,42^ (9%)) as outcome measures. Hemodynamics were evaluated in 14 reports (44%).^17,20–22,24,25,27,29–32,35,37,46^ Regarding hemodynamic features, various different endpoints were described: Aside from systolic, diastolic, and mean arterial blood pressure^10,20,21,27,30–32,35,37^, coronary perfusion pressure^17,32^, stroke volume,^17^ ejection fraction^17^, cardiac output^21,25^, coronary-^20^ or carotid flow^10,17,20,22,25,29^, aortic pressure and right atrial pressure^20–22,25,32^, central venous pressure,^21,30,35,37^ SpO2,^24^ as well as end tidal CO2 (etCO_2_)^17,24,27,32^ were assessed. Cerebral oxygenation (rSO_2_) was assessed in three studies^29,32,33^ (9%), and neuroinflammation was evaluated in another one^26^ (3%).

Patient outcomes were reported in nine studies (28%) ^3,15,23,29,32,38,40–42^, and adverse events, especially lung injuries, were discussed in six studies (19%).^3,21,25,27,31,39^ (Autopsy by predefined protocol – details not reported ^3^ / rib fractures, damage to the heart and lungs including abrasions, edema, ecchymosis, rupture of lung tissue^21^ / pulmonary lacerations, edema, bullae, hemorrhage or injury to thoracic cavity ^25^ / incidence of pneumothorax^27^ / diffuse alveolar hemorrhage, epithelia destruction, (hemo)pneumothorax^31^ / emphysema, intraparenchymal cysts, congestion, interstitial thickening, alveolar hemorrhage and exudates^39^)

### Various compression synchronized ventilation protocols

While 23 studies dealt with CCSV in particular ^15,19,26–35,37–43,47–50^, nine studies described different but mechanistically related compression synchronized ventilation algorithms^3,17,20–25,46^.

Ventilation simultaneous with chest compressions aside from CCSV was explored by seven studies (22%; three human^3,23,46^ and four animal studies^17,20–22^; airway pressure ranged from 20 cmH_2_O to 136 cmH_2_O). In one studies, synchronized ventilation was limited to a frequency of 10/minute. ^23^

*Schaller et al.* focused on automatic ventilation during decompression at a rate of 100/min in seven cases of OHCA in Munich, Germany ^24^, and *Olasveengen et al.* explored ventilations between chest compressions as well. ^25^

### Reported effects

Study populations and outcomes are summarized in *Table 2*.

- Clinical Outcomes: Beneficial effects of CCSV on cerebral TNFIZI mRNA levels in pigs were reported by *Renz et al.* ^26^ High rates of ROSC with CCSV and aortic ballon occlusion were observed in pigs by *Xu et al.* ^32^ The use of the mechanistically comparable ventilation mode “Cardiopulmonary Ventilation” (AIR LIQUIDE MEDICAL SYSTEMS SA, Antony, France; respiratory rate of 10/min, inspiratory pressure of 20 cmH_2_O and a PEEP of 5 cmH_2_O; magnification of intrathoracic pressure) was found to be associated with higher rates of ROSC in humans in a retrospective analysis of the Belgian cardiac arrest registry by *Malinverni et al.* ^23^ In contrast, *Krischer et al.* reported better survival rates to hospital admission and hospital discharge for conventional CPR compared with simultaneous compression and ventilation (at a rate of 40/min with an airway pressure of 80mmHg) in humans. ^3^ A recent, non-randomized study comparing CCSV, IPPV and bag ventilation during OHCA by *Hernández-Tejedor et al.* showed a non-significant trend towards higher rates of ROSC and favorable neurological survival (CPC 1/2 ) for patients ventilated with CCSV. ^42^
- Hemodynamics: *Chandra et al.* demonstrated improved carotid blood flow in animals and humans ^20,46^ and radial artery pressure in humans ^46^ during synchronized ventilation and compression when compared to conventional CPR. They also showed a correlation between lateral pleural pressure and aortic pressure and proposed it as a main determinant for blood flow during CPR in animals. ^22^ Similarly, *Kill et al.* reported improved mean arterial and coronary perfusion pressures with CCSV ^35^, and mean arterial pressures for CCSV in comparison to IPPV ^30,37^ or BiPAP ^30^ in pig models. Additionally, they were able to show improved cerebral oxygen saturation for CCSV when compared to IPPV.^33^ Similar results for cerebral oxygen saturation in pigs were reported by *Xu et al.* ^32^ and *Hu et al.* ^29^ The latter also demonstrated an improved carotid flow with CCSV when compared to IPPV, as well as lower lactate levels. ^29^ On the other hand *Kopra et al.* observed no effects of CCSV on arterial blood pressure in comparison to bag-valve ventilation also in a pig model.^27^ Only minimally improved hemodynamic effects of synchronized (between compressions) compared to unsynchronized manual ventilation at a rate of 10/min were observed by *Olasveengen et al.* also in a pig model. ^25^ *Hou et al.* showed an increase in central venous pressure in the simultaneous compression ventilation subgroup (at a ventilation rate of 60/min) when compared to standard CPR in pigs. ^21^
- Gas exchange: The majority of studies reported on gas exchange parameters^17,19–21,27,28,30–32,34,37,38,42^. *Kill et al.* showed adequate oxygenation as well as decarboxylation with CCSV during CPR ^34^, and a better results compared to IPPV and BiPAP.^30^ An association of higher paO_2_ with CCSV compared to IPPV was also described by *Renz et al., Xu et al., Kill et al. and Dersch et al.* ^31,32,37,38^ The same effect was observed by *Kopra et al.* in comparison to bag-valve ventilation.^27^ *Manegold et al.* described an additional benefit of the combination of aortic occlusion and CCSV regarding pO_2_, pCO_2_ and pH. ^28^ An improved paO_2_ was also described by *Cao et al.* in an Automatic Compression Synchronous Ventilation (ACSV; 3ml/kg tidal volume, respiratory rate 100/min) group compared to IPPV. ^17^ However, *Hou et al.* reported more a rapid deterioration of pH, paO_2_, and paCO_2_ in the simultaneous compression ventilation subgroup (at a rate of 60/min) compared to standard CPR. ^21^ Regarding deviations from ventilator presets (tidal volumes, peak airway pressures) CCSV outperformed IPPV and BiPAP in manikins as reported by *Speer et al.* ^43^ In humans *Schaller et al.* observed maximum etCO_2_ values below 44 mmHg during synchronized ventilation between compressions. ^24^ Only two, recently published studies examine the effects of CCSV on gas exchange in humans. ^15,42^ *Oh et al.* randomized OHCA patients at hospital arrival at a single center to a IPPV or CCSV ventilation group compared arterial pO_2_ and pCO_2_. Of 343 cases presenting at the hospital, 144 were randomized but only 30 patients (15 in each group) were successfully included with initial and follow-up blood gas analysis. They were able to show a significant increase in pO_2_ in the CCSV group and less marked, non-significant increase in the IPPV group, however, they failed to show a significant difference between ventilation groups (CCSV and IPPV). A significant decrease in pCO_2_ in the CCSV group was demonstrated.^15^ It has to be noted, that prehospital care of these OHCA patients was not standardized and not included in the analysis. In a non-randomized prospective study in OHCA patients (100 patients ventilated with CCSV, 145 ventilated with IPPV, and 276 with bag ventilation), *Hernández-Tejedor et al.* showed a significantly increased paO_2_ for patients ventilated with CCSV (7 blood samples) compared to IPPV (32 samples) and bag ventilation (24 samples). Arterial pH and paCO_2_ did not differ significantly. It has to be mentioned that arterial blood gas samples were not mandatory in this study. In venous blood samples, a non-significant trend towards higher pCO_2_ in patients with bag ventilation compared to mechanically ventilated patients was observed.^42^
- Adverse events: Data on adverse events are conflicting: *Kopra et al.* described a significantly higher prevalence of pneumothorax in the CCSV subgroup compared to bag-valve ventilation in pigs.^27^ *Renz et al.* observed higher epithelial damage associated with CCSV in pigs. ^31^ On the other hand, the rates of lung injuries in pigs were comparable between CCSV and IPPV as shown by *Dersch et al.* ^39^, and between synchronized and unsynchronized groups in the study by *Olasveengen et al.*^25^ *Kill et al*. reported no adverse events of CCSV in OHCA in humans. ^41^

**Table 2:**
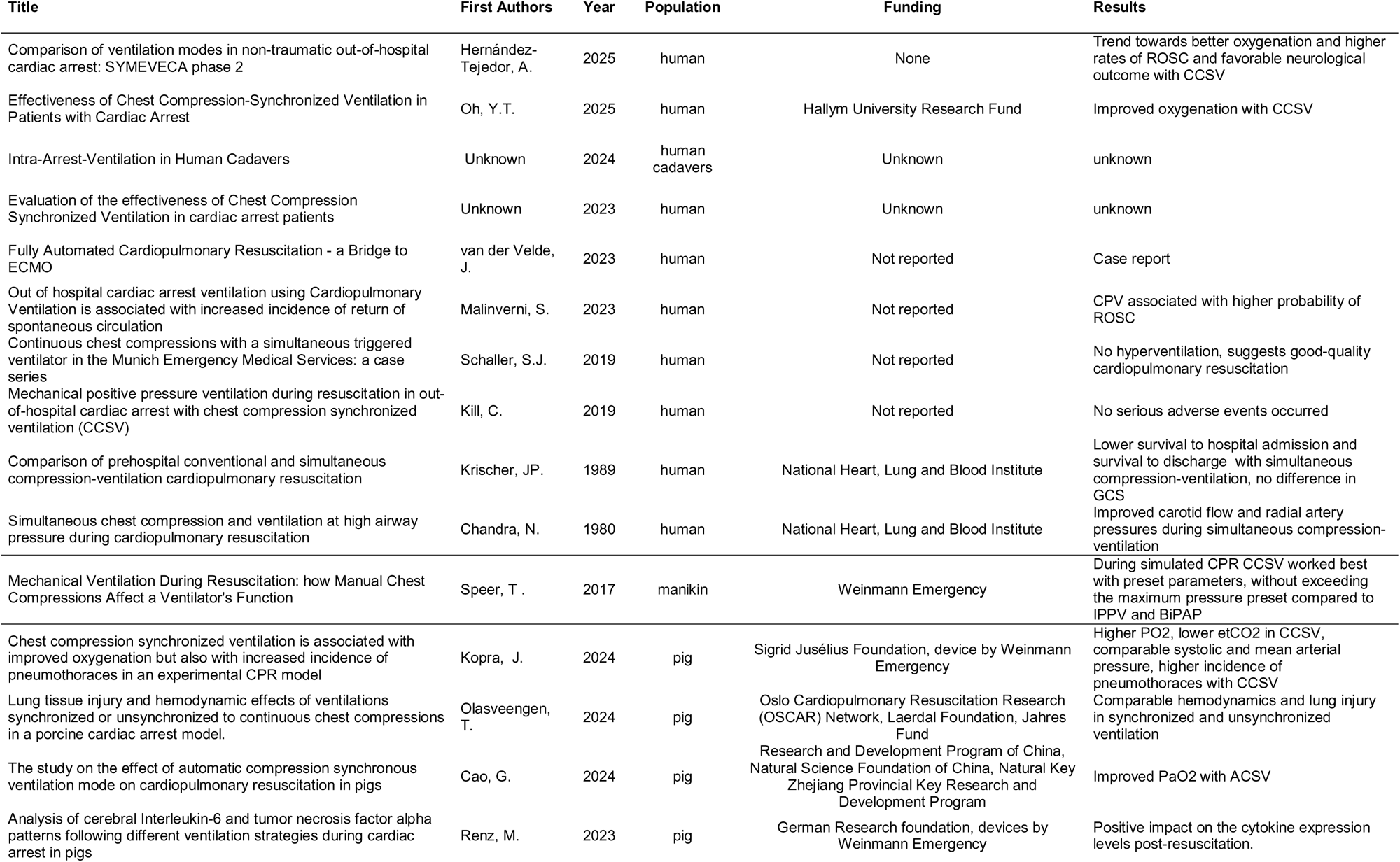
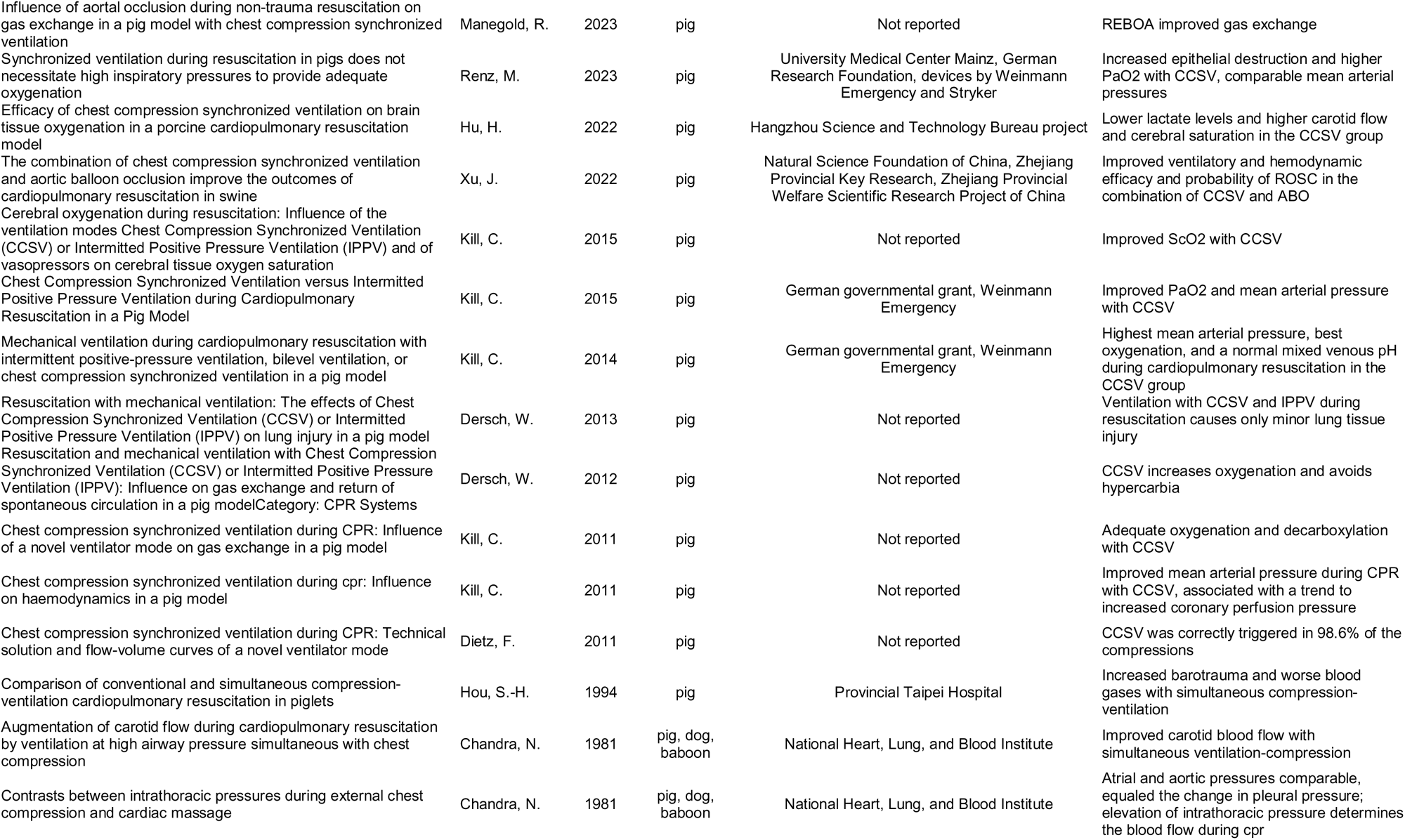
Overview of study populations and results, classified into animal and human studies and ordered by year of publication. (Missing authors due to missing full-text)

## Discussion

After reviewing numerous studies, it must be concluded that data on CCSV in humans are still scarce. On the other hand, a larger body of available evidence was identified for animals.

Oxygenation in terms of paO_2_ is generally described as superior for CCSV when compared to IPPV, BiPAP, or manual ventilation during cardiac arrest. ^30–32,34,37,38^ Beneficial effects of CCSV on oxygenation was also described in humans. ^15,42^ Despite the extremely high ventilation rate, an adequate decarboxylation avoiding hypercarbia is specifically suggested for CCSV. ^30,34^ Hemodynamic assessments during CPR are generally in favor of CCSV: beneficial effects on mean arterial-^30,35,37^ and coronary perfusion pressures^35^, as well as carotid flow and lower lactate concentrations^29^ have been described. Additionally, cerebral oxygenation as a combined endpoint for oxygenation and hemodynamics seems to be favorable during CCSV.^29,32,33^

However, despite these numerous physiological parameters, data on patient outcomes are scarce. Moreover, data on adverse events, especially lung injuries including pneumothorax due to the high ventilation pressures, are currently limited and conflicting, and may differ between animals and humans. Of note, no adverse events were described in an OHCA cohort of 34 human patients in Germany. ^41^ Additional safety data from larger prospective trials in humans are needed to clarify this.

It must also be mentioned that a large number of reports originate from Germany and the lion’s share of these from a research group of *Kill et al* (an overview of funding / sponsoring is given in *Table 2)*.

### Limitations

We found a considerable diversity of reported endpoints, especially hemodynamic outcomes, throughout the included studies. Additionally, the applied ventilation parameters varied largely, and some authors described and compared specific experimental ventilation protocols rather than standardized or commercialized ventilation modes. Therefore, comparability is limited and must be assessed cautiously. The importance of a common terminology regarding intra-arrest ventilation was also highlighted recently by *Segond et al.*^51^

With the intention to provide a broad overview of the current literature we included studies examining other protocols besides pure CCSV, e.g. CPV and SC-V, as well. Of course, direct comparability is not given in these cases and results have to be analyzed cautiously. Also, three full-texts could not be retrieved and therefore not assessed.

### Future outlook

A considerable proportion of studies focused on gas exchange or hemodynamic values which may be relevant physiological outcome measures serving as surrogate markers and are useful to understand mechanistic aspects of an intervention, but do not necessarily provide information about the effect of CCSV on survival outcomes. This is further aggravated by the scarcity of prospective clinical data on CCSV. Therefore, further clinical observational studies and randomized controlled trials reporting core outcome data along with rigorous safety monitoring are needed to provide reliable data on the effectiveness of CCSV during CPR. ^52^ Other aspects remain yet to be explored, for instance the impact of specialized ventilation modes such as CCSV on the management of human and team resources especially in a prehospital setting, and the general availability (and utilization) of specialized ventilation modes for CPR.

## Conclusion

CCSV is a novel and specialized approach for intra-arrest ventilation. Data from animal and human studies report improvements of physiological parameters during CPR but direct data on patient outcomes are still scarce. Prospective clinical trials in humans comparing the effectiveness of CCSV to other ventilation modes and manual ventilation are required.

## Declarations

### Data availability

The assessed data are available upon reasonable request to the corresponding author.

### Author contributions

CRediT author statement:

- Roman Brock: Conceptualization, Data curation, Investigation, Methodology, Validation, Visualization, Writing – original draft, Writing – review & editing
- Christoph Veigl: Data curation, Investigation, Writing – original draft, Writing – review & editing
- Andrea Kornfehl: Data curation, Investigation, Writing – review & editing
- Johannes Wittig: Validation, Writing – original draft, Writing – review & editing
- Sabine Heider: Data curation, Investigation, Writing – review & editing
- Karina Tapinova: Data curation, Investigation, Methodology, Writing – review & editing
- Erwin Snijders: Data curation, Investigation, Writing – review & editing
- Sabine Dunkl: Data curation, Investigation, Writing – review & editing
- Daniel Grassmann: Data curation, Investigation, Writing – review & editing
- Birgit Heller: Data curation, Investigation, Methodology, Resources, Validation, Writing – review & editing
- Mario Krammel: Data curation, Investigation, Writing – review & editing
- Sebastian Schnaubelt: Conceptualization, Data curation, Investigation, Methodology, Project administration, Resources, Supervision, Validation, Visualization, Writing – original draft, Writing – review & editing

## Acknowledgements

None.

