## Supplements for "Chest Compression Synchronized Ventilation (CCSV) during cardiac arrest in animals and humans: A scoping review"

### Search strategy for *Central*

| **#** | **Query** | **Results** |
| --- | --- | --- |
| 1 | "chest compression synchronized ventilat*".ti,ab,kw. | 3 |
| 2 | "chest compression synchronised ventilat*".ti,ab,kw. | 0 |
| 3 | CCSV.ti,ab,kw. | 5 |
| 4 | 1 or 2 or 3 | 6 |
| 5 | exp Cardiopulmonary Resuscitation/ | 1743 |
| 6 | ("Cardiopulmonary Resuscitation*" or "Cardio pulmonary Resuscitation*" or "cardiopulmonary reanimation*" or "cardio pulmonary reanimation*" or CPR or "basic life support*" or "advanced life support*" or "cardiac life support*" or "cardiovascular life support*" or "chest compression*" or "cardiac massage*" or "heart massage*" or "mouth to mouth Resuscitation*").ti,ab,kw. | 4671 |
| 7 | 5 or 6 | 5012 |
| 8 | 4 and 7 | 4 |
| 9 | exp Positive-Pressure Respiration/ | 3924 |
| 10 | ("positive pressure ventilat*" or "positive pressure respirat*").ti,ab,kw. | 2136 |
| 11 | ("pressure controlled ventilat*" or "pressure controlled respirat*").ti,ab,kw. | 443 |
| 12 | (simultaneous adj2 (ventilat* or respirat*)).ti,ab,kw. | 10 |
| 13 | ("synchronized ventilat*" or "synchronized respirat*").ti,ab,kw. | 19 |
| 14 | ((new or novel) adj3 (ventilat* or respirat*)).ti,ab,kw. | 564 |
| 15 | 9 or 10 or 11 or 12 or 13 or 14 | 6355 |
| 16 | 7 and 15 | 118 |
| 17 | exp Heart Arrest/ | 3068 |
| 18 | ("heart arrest*" or "cardiac arrest*" or Asystol* or "Cardiopulmonary Arrest*" or "cardio pulmonary arrest*" or "circulation arrest*" or "circulatory arrest*" or "heart standstill*").ti,ab,kw. | 6087 |
| 19 | 17 or 18 | 7223 |
| 20 | 16 and 19 | 37 |
| 21 | 8 or 20 | 39 |

Supplemental Table 1: Search strategy for Central

### Search Strategy for *Embase*

| **No.** | **Query** | **Results** |
| --- | --- | --- |
| #22 | #8 OR #21 | 571 |
| #21 | #17 AND #20 | 564 |
| #20 | #18 OR #19 | 165702 |
| #19 | 'heart arrest*':ti,ab,kw OR 'cardiac arrest*':ti,ab,kw OR asystol*:ti,ab,kw OR 'cardiopulmonary arrest*':ti,ab,kw OR 'cardio pulmonary arrest*':ti,ab,kw OR 'circulation arrest*':ti,ab,kw OR 'circulatory arrest*':ti,ab,kw OR 'heart standstill*':ti,ab,kw | 100026 |
| #18 | 'heart arrest'/exp | 147187 |
| #17 | #7 AND #16 | 1152 |
| #16 | #9 OR #10 OR #11 OR #12 OR #13 OR #14 OR #15 | 93867 |
| #15 | ((new OR novel) NEAR/3 (ventilat* OR respirat*)):ti,ab,kw | 6999 |
| #14 | 'synchronized ventilat*':ti,ab,kw OR 'synchronized respirat*':ti,ab,kw | 128 |
| #13 | (simultaneous NEAR/2 (ventilat* OR respirat*)):ti,ab,kw | 311 |
| #12 | 'pressure controlled ventilat*':ti,ab,kw OR 'pressure controlled respirat*':ti,ab,kw | 1012 |
| #11 | 'positive pressure ventilat*':ti,ab,kw OR 'positive pressure respirat*':ti,ab,kw | 11552 |
| #10 | 'pressure controlled ventilation'/exp | 842 |
| #9 | 'positive pressure ventilation'/exp | 84013 |
| #8 | #4 AND #7 | 20 |
| #7 | #5 OR #6 | 57879 |
| #6 | 'cardiopulmonary resuscitation*':ti,ab,kw OR 'cardio pulmonary resuscitation*':ti,ab,kw OR 'cardiopulmonary reanimation*':ti,ab,kw OR 'cardio pulmonary reanimation*':ti,ab,kw OR cpr:ti,ab,kw OR 'basic life support*':ti,ab,kw OR 'advanced life support*':ti,ab,kw OR 'cardiac life support*':ti,ab,kw OR 'cardiovascular life support*':ti,ab,kw OR 'chest compression*':ti,ab,kw OR 'cardiac massage*':ti,ab,kw OR 'heart massage*':ti,ab,kw OR 'mouth to mouth resuscitation*':ti,ab,kw | 57495 |
| #5 | 'advanced cardiac life support'/de | 901 |
| #4 | #1 OR #2 OR #3 | 39 |
| #3 | ccsv:ti,ab,kw | 37 |
| #2 | 'chest compression synchronised ventilat*':ti,ab,kw | 1 |
| #1 | 'chest compression synchronized ventilat*':ti,ab,kw | 18 |

Supplemental Table 2: Search strategy for Embase

### Search Strategy for *Medline*

| **#** | **Query** | **Results** |
| --- | --- | --- |
| 1 | "chest compression synchronized ventilat*".ti,ab,kf. | 8 |
| 2 | "chest compression synchronised ventilat*".ti,ab,kf. | 0 |
| 3 | CCSV.ti,ab,kf. | 26 |
| 4 | 1 or 2 or 3 | 28 |
| 5 | exp Cardiopulmonary Resuscitation/ | 23914 |
| 6 | ("Cardiopulmonary Resuscitation*" or "Cardio pulmonary Resuscitation*" or "cardiopulmonary reanimation*" or "cardio pulmonary reanimation*" or CPR or "basic life support*" or "advanced life support*" or "cardiac life support*" or "cardiovascular life support*" or "chest compression*" or "cardiac massage*" or "heart massage*" or "mouth to mouth Resuscitation*").ti,ab,kf. | 36730 |
| 7 | 5 or 6 | 45269 |
| 8 | 4 and 7 | 8 |
| 9 | exp Positive-Pressure Respiration/ | 30316 |
| 10 | ("positive pressure ventilat*" or "positive pressure respirat*").ti,ab,kf. | 7882 |
| 11 | ("pressure controlled ventilat*" or "pressure controlled respirat*").ti,ab,kf. | 719 |
| 12 | (simultaneous adj2 (ventilat* or respirat*)).ti,ab,kf. | 237 |
| 13 | ("synchronized ventilat*" or "synchronized respirat*").ti,ab,kf. | 89 |
| 14 | ((new or novel) adj3 (ventilat* or respirat*)).ti,ab,kf. | 5234 |
| 15 | 9 or 10 or 11 or 12 or 13 or 14 | 39638 |
| 16 | 7 and 15 | 519 |
| 17 | exp Heart Arrest/ | 59233 |
| 18 | ("heart arrest*" or "cardiac arrest*" or Asystol* or "Cardiopulmonary Arrest*" or "cardio pulmonary arrest*" or "circulation arrest*" or "circulatory arrest*" or "heart standstill*").ti,ab,kf. | 63190 |
| 19 | 17 or 18 | 91110 |
| 20 | 16 and 19 | 217 |
| 21 | 8 or 20 | 219 |

Supplemental Table 3: Search strategy for Medline

### Search Strategy for *Scopus*

| **#** | **Query** | **Results** |
| --- | --- | --- |
| 1 | TITLE-ABS-KEY ( ( "chest compression synchronized ventilat*" ) ) | 9 |
| 2 | TITLE-ABS-KEY ( ( "chest compression synchronised ventilat*" ) ) | 9 |
| 3 | TITLE-ABS-KEY ( ( ccsv ) ) | 56 |
| 4 | 1 OR 2 OR 3 | 59 |
| 5 | TITLE-ABS-KEY ( ( "Cardiopulmonary Resuscitation*" OR "Cardio pulmonary Resuscitation*" OR "cardiopulmonary reanimation*" OR "cardio pulmonary reanimation*" OR cpr OR "basic life support*" OR "advanced life support*" OR "cardiac life support*" OR "cardiovascular life support*" OR "chest compression*" OR "cardiac massage*" OR "heart massage*" OR "mouth to mouth Resuscitation*" ) ) | 58954 |
| 6 | 4 AND 5 | 9 |
| 7 | TITLE-ABS-KEY ( ( "positive pressure ventilat*" OR "positive pressure respirat*" ) ) | 27576 |
| 8 | TITLE-ABS-KEY ( ( "pressure controlled ventilat*" OR "pressure controlled respirat*" ) ) | 1331 |
| 9 | TITLE-ABS-KEY ( ( ( simultaneous ) near/2 ( ventilat* OR respirat* ) ) ) | 137 |
| 10 | TITLE-ABS-KEY ( ( "synchronized ventilat*" OR "synchronized respirat*" ) ) | 125 |
| 11 | TITLE-ABS-KEY ( ( ( new OR novel ) near/3 ( ventilat* OR respirat* ) ) ) | 887 |
| 12 | 7 OR 8 OR 9 OR 10 OR 11 | 29692 |
| 13 | 5 AND 12 | 641 |
| 14 | TITLE-ABS-KEY ( ( "heart arrest*" OR "cardiac arrest*" OR asystol* OR "Cardiopulmonary Arrest*" OR "cardio pulmonary arrest*" OR "circulation arrest*" OR "circulatory arrest*" OR "heart standstill*" ) ) | 120603 |
| 15 | 13 AND 14 | 287 |
| 16 | 6 OR 15 | 289 |

Supplemental Table 4: Search strategy for Scopus

### Search Strategy for *Web of Science*

| **#** | **Search Query** | **Results** |
| --- | --- | --- |
| 1 | TS=("chest compression synchronized ventilat*") | 10 |
| 2 | TS=("chest compression synchronised ventilat*") | 0 |
| 3 | TS=(CCSV) | 43 |
| 4 | #1 OR #2 OR #3 | 46 |
| 5 | TS=("Cardiopulmonary Resuscitation*" or "Cardio pulmonary Resuscitation*" or "cardiopulmonary reanimation*" or "cardio pulmonary reanimation*" or CPR or "basic life support*" or "advanced life support*" or "cardiac life support*" or "cardiovascular life support*" or "chest compression*" or "cardiac massage*" or "heart massage*" or "mouth to mouth Resuscitation*") | 47494 |
| 6 | #4 AND #5 | 10 |
| 7 | TS=("positive pressure ventilat*" or "positive pressure respirat*") | 9905 |
| 8 | TS=("pressure controlled ventilat*" or "pressure controlled respirat*") | 765 |
| 9 | TS=((simultaneous) NEAR/2 (ventilat* or respirat*)) | 623 |
| 10 | TS=("synchronized ventilat*" or "synchronized respirat*") | 91 |
| 11 | TS=((new or novel) NEAR/3 (ventilat* or respirat*)) | 8986 |
| 12 | #7 OR #8 OR #10 OR #9 OR #11 | 20071 |
| 13 | #5 AND #12 | 485 |
| 14 | TS=("heart arrest*" or "cardiac arrest*" or Asystol* or "Cardiopulmonary Arrest*" or "cardio pulmonary arrest*" or "circulation arrest*" or "circulatory arrest*" or "heart standstill*") | 76050 |
| 15 | #13 AND #14 | 172 |
| 16 | #15 OR #6 | 174 |

Supplemental Table 5: Search strategy for Web of Science
